## Supplementary Material for "When should lockdown be implemented? Devising cost-effective strategies for managing epidemics amid vaccine uncertainty"

### 1 Model Description

Here we present a detailed description of the model used to simulate disease dynamics as briefly outlined in the main text. The population size is $N = 1$ million, disaggregated by three demographic cohorts $G = \{0-19,\ 20-64,\ 65+\}$ of sizes $N_{0-19} = 233,000$, $N_{20-64} = 580,000$, $N_{65+} = 187,000$ to resemble a subset of the United Kingdom demography structure [1].

Individuals who are susceptible ($S_a$) can acquire infection from infectious individuals and become exposed ($E_{a,m}$) before progressing to the infectious compartments upon the completion of the latent period. The model distinguishes between four infectious compartments per age cohort. Infectious individuals can have either symptomatic infection with age-dependent probability $d_a$ or asymptomatic ($I^A_{a,m}$) infection without developing symptoms with probability $1 - d_a$. Symptomatic infection is further subdivided into compartments for individuals who are symptomatic without requiring hospital treatment ($I^S_{a,m}$) with age-dependent probability $d_a(1 - h_a)$, symptomatic who will inevitably require hospital treatment ($I^{PH}_{a,m}$) with probability $d_a h_a$ or individuals in hospital with severe disease ($I^H_a$), who have progressed from the "pre-hospitalisation" compartment ($I^{PH}_{a,m}$) at rate $\gamma$. All infectious classes contribute towards pathogen transmission, with reduced levels of infectiousness $\tau$ assumed for asymptomatic individuals in comparison to symptomatic individuals. Additionally, it is assumed that individuals in the pre-hospital compartment carry the same contact rate as individuals with general symptomatic infection, but hospitalised individuals are less infectious by a factor $\rho$ due to ward isolation. Individuals in hospital (who avoid disease-induced mortality with age-dependent probability $1 - \mu_a$) recover at a reduced rate $\delta < \gamma$. The final consideration to the model structure is to incorporate waning immunity and the possibility of reinfection. Hence, a recovered class $R_{a,m}$ is modelled explicitly where individuals progress to upon the completion of the infectious stages. Recovered individuals remain in the class until immunity wanes at rates $\omega$ and they return to susceptibility, this rate is chosen to be sufficiently small as to not overestimate the number of reinfections in the early stages of the outbreak. It should be noted that once individuals return to the susceptible compartment, the probability of severe disease is independent of previous infection status.

We apply the method of stages to ensure that the latent, infectious and recovered periods are gamma distributed as opposed to exponentially distributed in the equivalent stochastic framework. To enforce a latent period with a total length $\epsilon^{-1}$ which is approximately Erlang distributed, there are $M = 3$ exposed compartments ($E_{a,1}$, $E_{a,2}$, $E_{a,3}$) for individuals who have been infected but are not yet infectious. Similarly, there are $M$ compartments each for asymptomatic infection, symptomatic infection and recovery ($I^A_{a,m}$, $I^S_{a,m}$, $I^{PH}_{a,m}$, $R_{a,m}$ with $m \in \{1, 2, 3\}$) such that the rates for leaving these classes $\gamma^{-1}$ and $\omega^{-1}$ are Erlang distributed with shape $M = 3$ in the equivalent stochastic framework. The exception is the hospitalisation stage which is constrained to a single compartment $I^H_a$. The length of the hospitalisation period $\delta^{-1}$ would be exponentially distributed if modelled explicitly. The length of stay in hospital follows a distribution which is approximately exponential and the choice of $\delta$ parameter was decided by fitting an exponential curve to this distribution. Due to the short simulation timescales of this study (on the order of 2-3 years) and the broad size of the age bins, the model does not account for natural births, deaths and ageing between demographic cohorts. The model dynamics are governed by a set of coupled ordinary differential equations (ODEs) for each demographic cohort $a \in G$, for $M = 3$ and $m \in \{2, 3\}$:

$$\begin{aligned}
\frac{dS_a}{dt} &= -\frac{S_a\lambda_a}{N_a} + M\omega R_{a,M},\\
\frac{dE_{a,1}}{dt} &= \frac{S_a\lambda_a}{N_a} - M\epsilon E_{a,1},\\
\frac{dE_{a,m}}{dt} &= M\epsilon(E_{a,m-1} - E_{a,m}),\\
\frac{dI^A_{a,1}}{dt} &= M\epsilon(1-d_a)E_{a,M} - M\gamma I^A_{a,1},\\
\frac{dI^A_{a,m}}{dt} &= M\gamma(I^A_{a,m-1} - I^A_{a,m}),\\
\frac{dI^S_{a,1}}{dt} &= M\epsilon d_a(1-h_a)E_{a,M} - M\gamma I^S_{a,1},\\
\frac{dI^S_{a,m}}{dt} &= M\gamma(I^S_{a,m-1} - I^S_{a,m}),\\
\frac{dI^{PH}_{a,1}}{dt} &= M\epsilon d_a h_a E_{a,M} - M\gamma I^{PH}_{a,1},\\
\frac{dI^{PH}_{a,m}}{dt} &= M\gamma(I^{PH}_{a,m-1} - I^{PH}_{a,m}),\\
\frac{dI^H_a}{dt} &= M\gamma I^{PH}_{a,M} - \delta I^H_a,\\
\frac{dR_{a,1}}{dt} &= M\gamma(I^A_{a,M} + I^S_{a,M}) + (1-\mu_a)\delta I^H_a - M\omega R_{a,1},\\
\frac{dR_{a,m}}{dt} &= M\omega(R_{a,m-1} - R_{a,m}).
\end{aligned} \tag{1}$$

Transmission occurs in accordance to the age-specific force of infection,

$$\lambda_a = \sum_{b\in G}\beta_{ba}\Big(\sum_{m\in\{1,2,3\}}(\tau I^A_{b,m} + I^S_{b,m} + I^{PH}_{b,m}) + \rho I^H_b\Big). \tag{2}$$

The parameter $\beta_{ba}$ captures transmission from individuals in cohort $b$ to individuals in cohort $a$. Transmission is driven by an age-structured contact matrix $\pi$ for the United Kingdom taken from Prem *et al.* [2]. Each entry $\pi_{ij}$ is an aggregation of the number of contacts with age group $j$ recorded by age group $i$.

$$\pi = \begin{pmatrix} 6.8045 & 4.0791 & 0.2121\\ 1.9898 & 8.1466 & 0.55565\\ 0.5221 & 3.4354 & 2.0034 \end{pmatrix}. \tag{3}$$

This matrix is scaled by a transmission factor $\Pi = \Pi(R_0)$ in order to obtain a transmission matrix $\beta$ such that the basic reproduction number will be $R_0 = 3.0$ upon numerical evaluation using the Next Generation Matrix (NGM) approach [3]. An interpretation of the transmission factor is that $1/\Pi$ is the probability of a transmission event occurring given contact. The remaining model parameters are fixed and their description and values are provided in Table S1.

$$\beta_{ij} = \frac{1}{\Pi}\pi_{ij}. \tag{4}$$

The implementation of control is modelled by adjusting all contact rates by the intensity of the control state $u(t)$ – equivalent to the relative reduction in transmission. This gives time-dependent contact rates,

$$\beta_{ij}(t) = (1 - u(t))\beta_{ij}. \tag{5}$$

The age-specific parameters $d_a$, $h_a$, $\mu_a$ in addition to the recovery rate from hospitalisation $\delta$ were originally modelled as distributions by Keeling *et al.* [4], which are publicly available. To apply the age-specific distributions to this study, they were aggregated from the original five-year age bands $G' = \{0{-}4, 5{-}9, \ldots, 95{-}100, 100+\}$ to match the three age cohorts $G$ used in this study. By defining subsets of the relevant age bands $G'_{0-19} = \{0-4, \ldots, 15-19\}$, $G'_{20-64} = \{20-24, \ldots, 60-64\}$, $G'_{65+} = \{65-69, \ldots, 100+\}$, the probability of symptomatic infection $d_a$ is computed as follows,

$$d_a = \sum_{g \in G'_a} N_{g,a} d'_g\,, \qquad a \in G, \tag{6}$$

where $N_{g,a}$ is the proportion of the population group $a$ which reside in age group $g$ (taken from [1]) and $d'_g$ is the original distribution for age bands $G'$ [4]. The same methodology is used to compute $h_a$, $\mu_a$. The time spent in the hospitalised class ($X$, measured in days) is assumed to obey an exponential distribution with mean $1/\delta$. The rate for leaving hospital $\delta$ is inferred by fitting the cumulative distribution function $F_X(x) = 1 - e^{-\delta x}$ to the cumulative distribution time in hospital distribution from Keeling *et al.* [4] using nonlinear least squares.

| Parameter | Description | Estimate | Units | Source |
|---|---|---|---|---|
| $R_0$ | Basic reproduction number | 3.0 | N/A | Assumed |
| $\Pi$ | Transmission scaling for $\beta$ | 17.9241 | N/A | Computed |
| $\tau$ | Relative asymptomatic infectiousness | 0.25 | N/A | [4] |
| $\rho$ | Relative infectiousness of hospitalised individuals | 0.1 | N/A | Assumed |
| $\epsilon$ | Latency progression rate | 1/5.28 | days$^{-1}$ | [5] |
| $d_{0-19}$ | Probability of developing symptoms (age 0-19) | 0.0275 | N/A | [4] |
| $d_{20-64}$ | Probability of developing symptoms (age 20-64) | 0.1350 | N/A | [4] |
| $d_{65+}$ | Probability of developing symptoms (age 65+) | 0.5374 | N/A | [4] |
| $h_{0-19}$ | Probability of hospitalisation given symptoms (age 0-19) | 0.1268 | N/A | [4] |
| $h_{20-64}$ | Probability of hospitalisation given symptoms (age 20-64) | 0.1173 | N/A | [4] |
| $h_{65+}$ | Probability of hospitalisation given symptoms (age 65+) | 0.2430 | N/A | [4] |
| $\mu_{0-19}$ | Probability of death given hospitalisation (age 0-19) | 0.0276 | N/A | [4] |
| $\mu_{20-64}$ | Probability of death given hospitalisation (age 20-64) | 0.0533 | N/A | [4] |
| $\mu_{65+}$ | Probability of death given hospitalisation (age 65+) | 0.1935 | N/A | [4] |
| $\gamma$ | Recovery rate from infection (outside of hospital) | 1/5 | days$^{-1}$ | [6] |
| $\delta$ | Recovery rate from hospitalisation | 1/8.78 | days$^{-1}$ | [4] |
| $\omega$ | Waning immunity rate | 1/800 | days$^{-1}$ | Assumed |

**Table S1: Disease parameters.** Table of fixed parameters and their biological interpretation in the mathematical model (1). Sensitivity to assumed parameters $R_0, \rho$ and $\omega$ are presented in Figures S12, S13, S16.

#### 2 Vaccination uncertainty

A total of four joint probability distributions were used to explore distributions in cost for each control strategy as the arrival date $T$ and eventual coverage $\eta$ of the vaccine varied. These were constructed by combining pairs of two marginal distributions each for $T$ and $\eta$:

$$\begin{aligned} T &= 60 + 60Y, \\ 1080 - T &= 60Y. \\ \eta &= 0.05Z, \\ 1 - \eta &= 0.05Z, \end{aligned} \tag{7}$$

where $Y$ and $Z$ are random variables defined over a sequence of integers $y = 0, 1, \ldots, 17$ and $z = 0, 1, \ldots, 20$ respectively. Their probability mass functions

$$\begin{aligned} \mathbb{P}[Y = y] &= e^{-k}\frac{k^y}{y!}/\Omega_Y, \\ \mathbb{P}[Z = z] &= e^{-l}\frac{l^z}{z!}/\Omega_Z, \end{aligned} \tag{8}$$

are analogous to the Poisson distribution with rates $k = 6$ and $l = 5$, subject to normalising constants $\Omega_Y, \Omega_Z$ to ensure that $\sum_y \mathbb{P}[Y = y] = 1 = \sum_z \mathbb{P}[Z = z]$. As showcased in the Figure S1, the Poisson distribution allows for sufficient variance across $T, \eta$ outcomes for the marginal distributions while maintaining very low probabilities at the extremities $\mathbb{P}[T] = \{60, 1080\}$, $\mathbb{P}[\eta] = \{0, 1\}$ of the relevant sample space. The final joint probability distributions arising from these marginal distributions are displayed in Figure S2.

#### 3 Additional figures

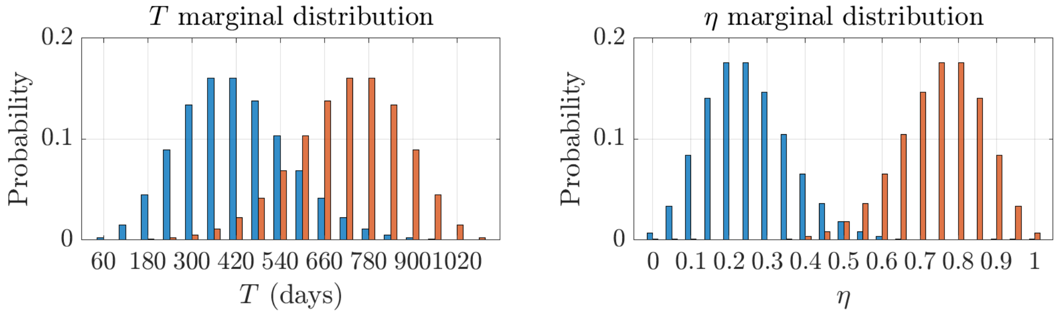


**Figure S1: Vaccine distributions (marginal).** Marginal probability distributions for the vaccine deployment date $T$ (left) and eventual coverage $\eta$ (right). The distributions are generated using the random variables $Y, Z$ who arise from discrete normal distributions with respective parameterisation $(\mu_T, \sigma_T^2)$ and $(\mu_\eta, \sigma_\eta^2)$. Different expectations for $T, \eta$ are driven by varying $\mu_T, \mu_\eta$ and the underlying variance is fixed, $\sigma_T^2 = \sigma_\eta^2 = 2.4$.

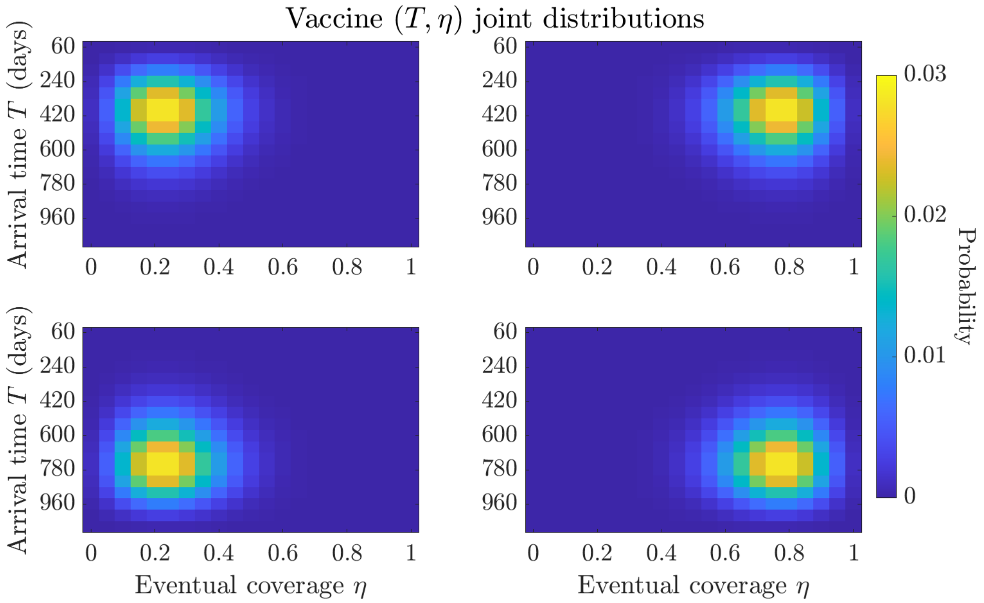


**Figure S2: Vaccine distributions (joint).** Joint probability distributions for vaccination uncertainty characterised by deployment date $T$ and eventual coverage $\eta$. The joint distributions cover a range of optimistic (top right: low $T$ and high $\eta$) and pessimistic (bottom left: high $T$ and low $\eta$) scenarios regarding the effectiveness of the vaccination campaign.

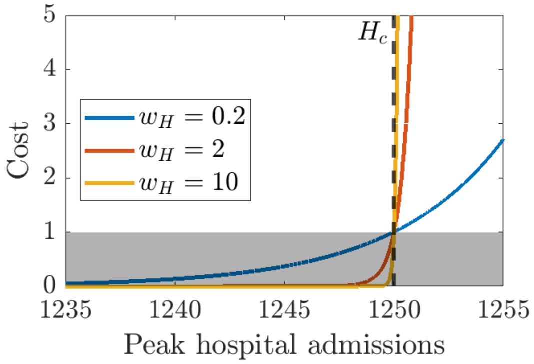


**Figure S3: Qualitative behaviour of exponential term in the objective function.** Qualitative behaviour of the exponential term in the objective function (1). Since the public health and control contributions are bounded in [0,1] (shaded region), any excess on peak hospital capacity $H_c$ renders the strategy infeasible with a cost exceeding at least one, regardless of weight $w_H$. The weight $w_H$ can be scaled to deal with different perspectives regarding risk to overwhelm, but will be fixed at $w_H = 2$ (red) for this study.

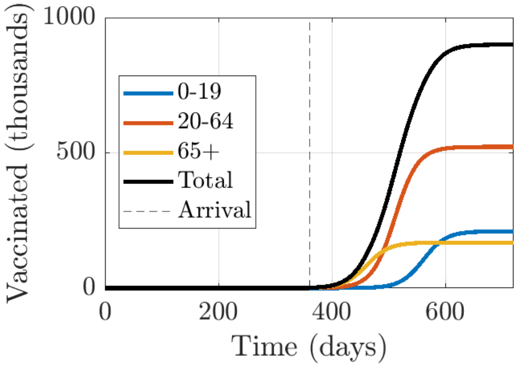


**Figure S4: Qualitative behaviour of vaccination parameters.** Cumulative vaccinations by age cohort $V_a(t)$ in a chosen simulation with parameter choices $\eta = 0.9, \kappa = 0.05, t_c = 100$ days and deployment date $T = 360$ days.

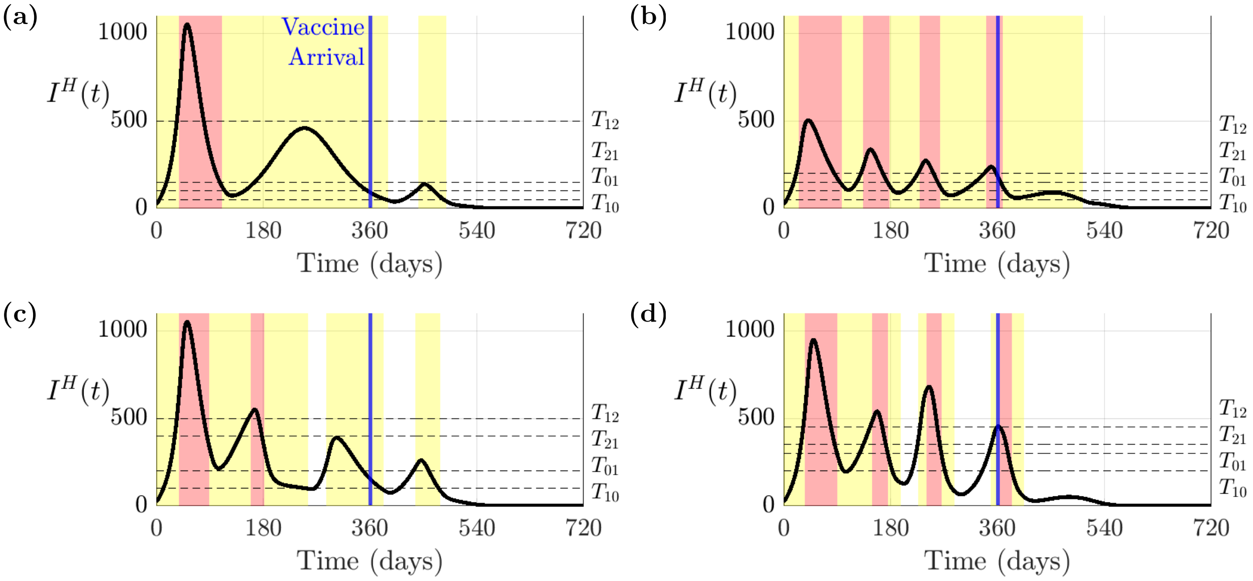


**Figure S5: Hospitalisation dynamics with vaccination.** Number of active hospitalised individuals across all age cohorts $I^H(t)$ for each control strategy, with a vaccine introduced on day $T = 360$ with $\eta = 0.9$. Strategies are labelled as (a) S1 (Cautious Easing), (b) S2 (Suppression), (c) S3 (Slow Control) and (d) S4 (Rapid Control). The periods spent in the Lockdown state and the Intermediate Control state are shaded in red and yellow respectively. The switching thresholds $T_{ij}$ are marked with horizontal dashed lines.

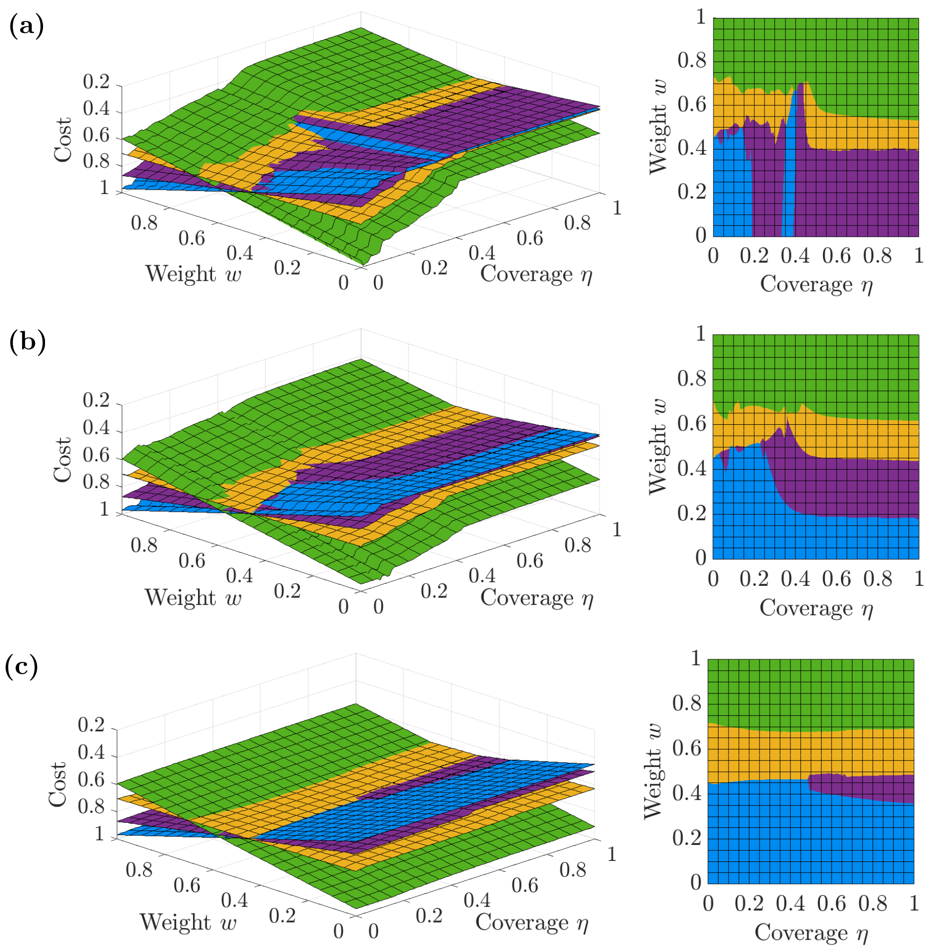


**Figure S6: Outbreak costs under different control strategies for different assumptions about the timing of vaccine development and the eventual vaccination coverage.** Surface plots of cost (evaluated by the objective function (1)) for different values of the weighting $w$ and the eventual coverage of the vaccine (left). The z-axis has the lowest cost at the top, allowing the optimal strategy (corresponding to the lowest cost) to be seen more clearly. Results were generated separately for vaccine time to deployment (a) $T = 360$, (b) $T = 630$ and (c) $T = 900$ days. The control strategies are coloured as follows: yellow (Cautious easing), green (Suppression), purple (Slow control) and blue (Rapid control).

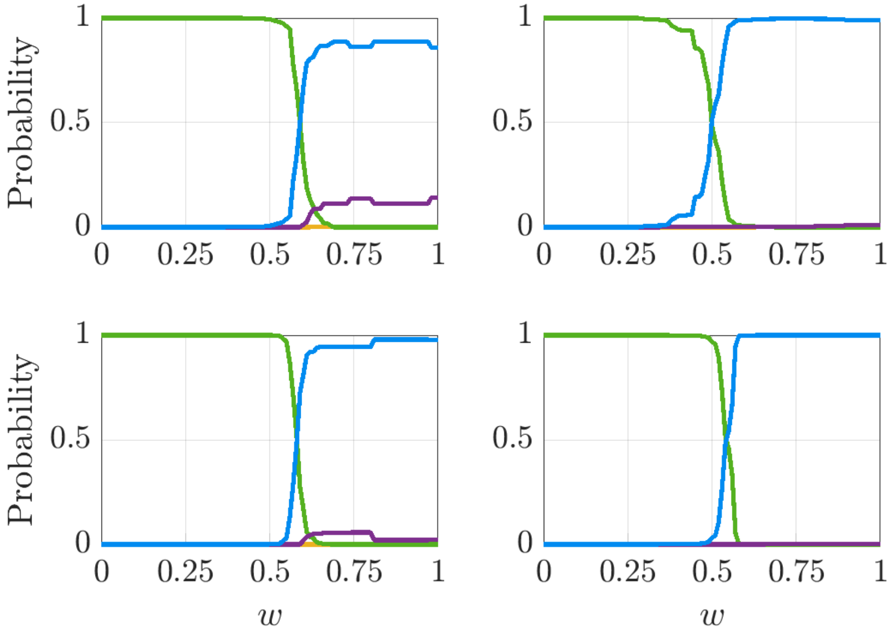


**Figure S7: Probability each control strategy accurs the greatest cost, subject to underlying vaccine distribution.** Probability that each control strategy has the greatest cost according to underlying vaccine joint distribution and weight $w_1$. The joint distributions cover a range of optimistic (top right: low $T$ and high $\eta$) and pessimistic (bottom left: high $T$ and low $\eta$) scenarios regarding the effectiveness of the vaccination campaign. This probability is measured by ranking the strategies across all permutations of timing and coverage parameters $(T, \eta)$ and summing the probabilities where each strategy is the extremum. The control strategies are coloured as follows: yellow (Cautious easing), green (Suppression), purple (Slow control) and blue (Rapid control).

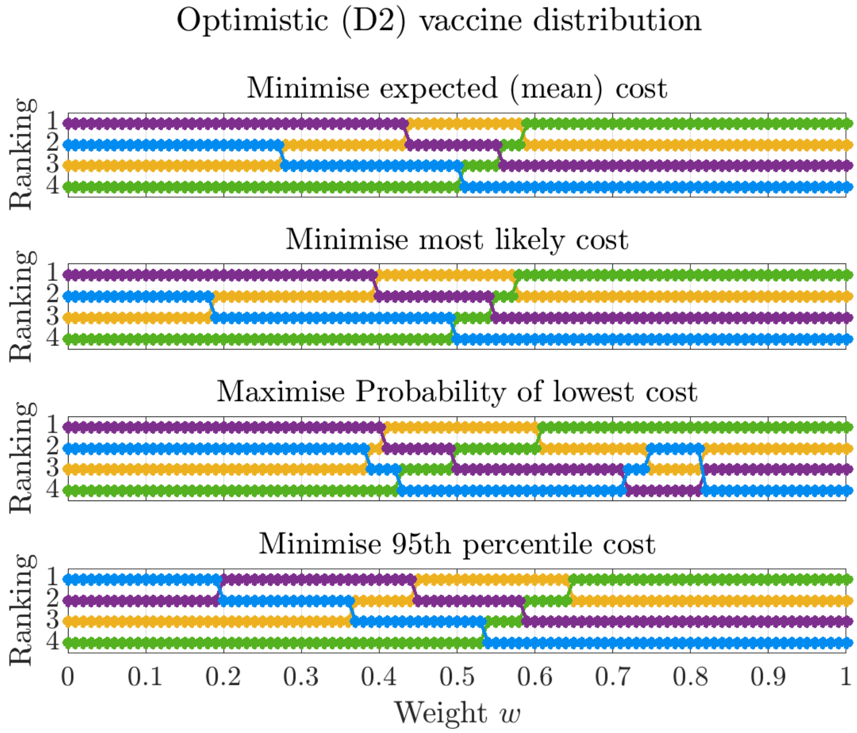


**Figure S8: Varying control strategy rankings by criteria.** Control strategy rankings according to a range of criteria described in Methods. Given a weighting $w_1$, the optimal strategy is sensitive to the summary statistic for which the policy maker seeks to minimise the objective function against. The control strategies are coloured as follows: yellow (Cautious easing), green (Suppression), purple (Slow control) and blue (Rapid control).

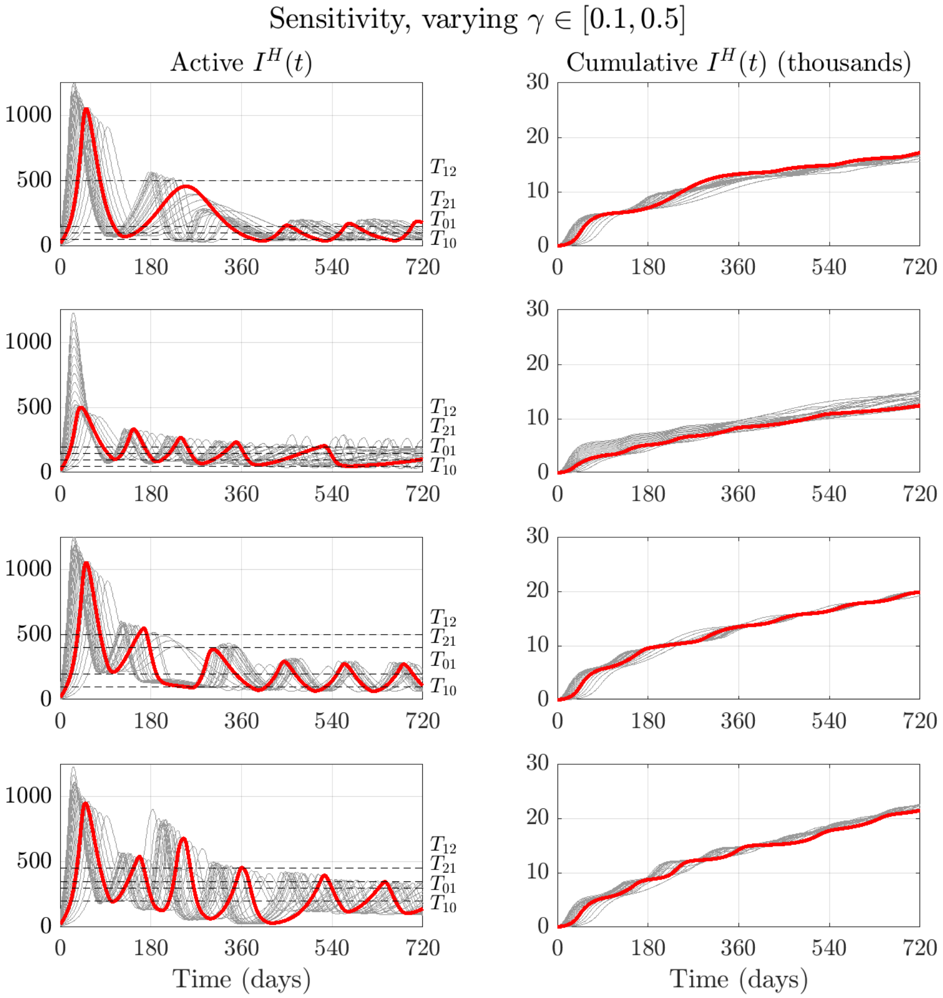


**Figure S9: Sensitivity to biological parameters.** Sensitivity in simulation dynamics to varying select parameters $\gamma$ while keeping the remaining parameters fixed.

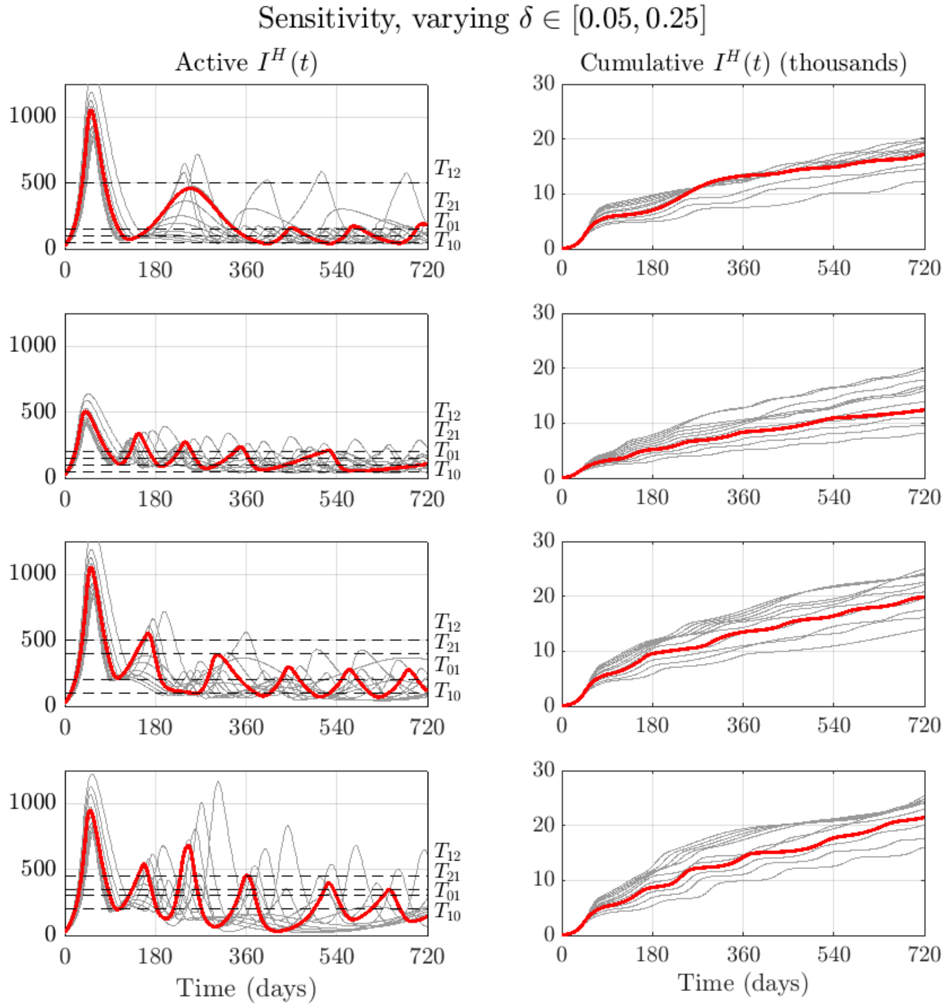


**Figure S10: Sensitivity to biological parameters.** Sensitivity in simulation dynamics to varying select parameters $\delta$ while keeping the remaining parameters fixed.

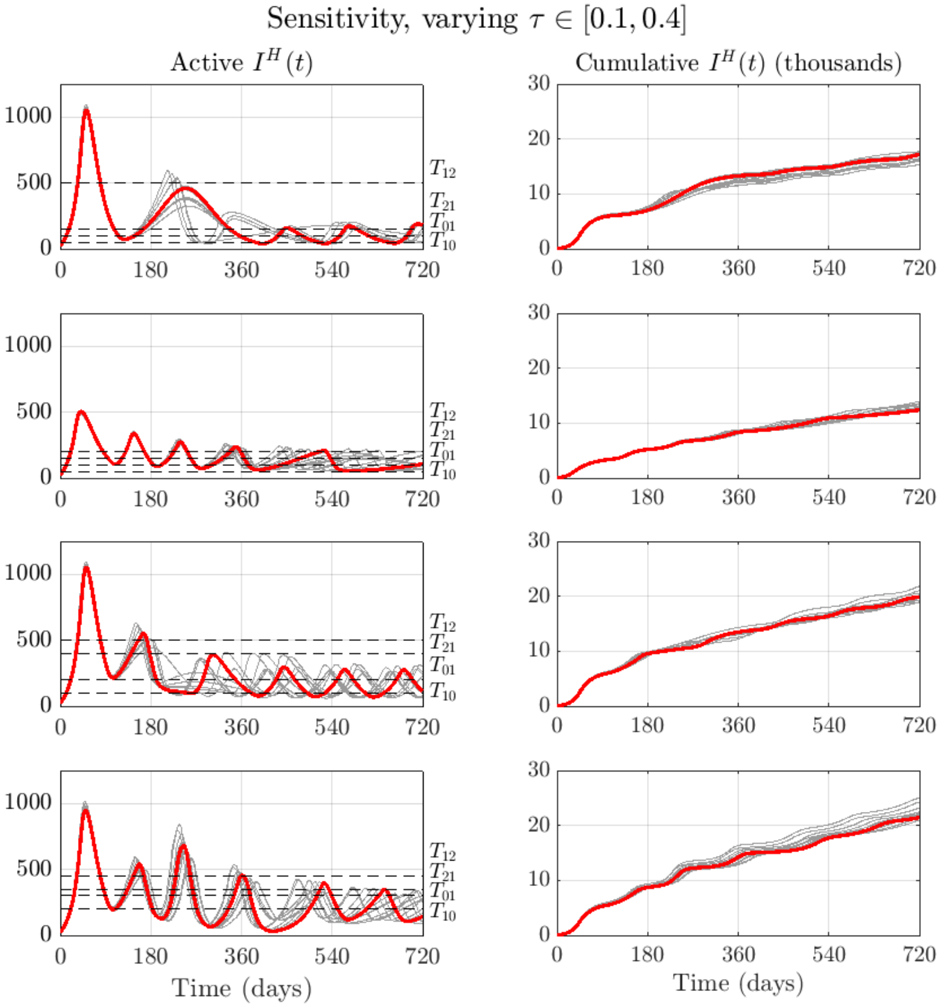


**Figure S11: Sensitivity to biological parameters.** Sensitivity in simulation dynamics to varying select parameters $\tau$ while keeping the remaining parameters fixed.

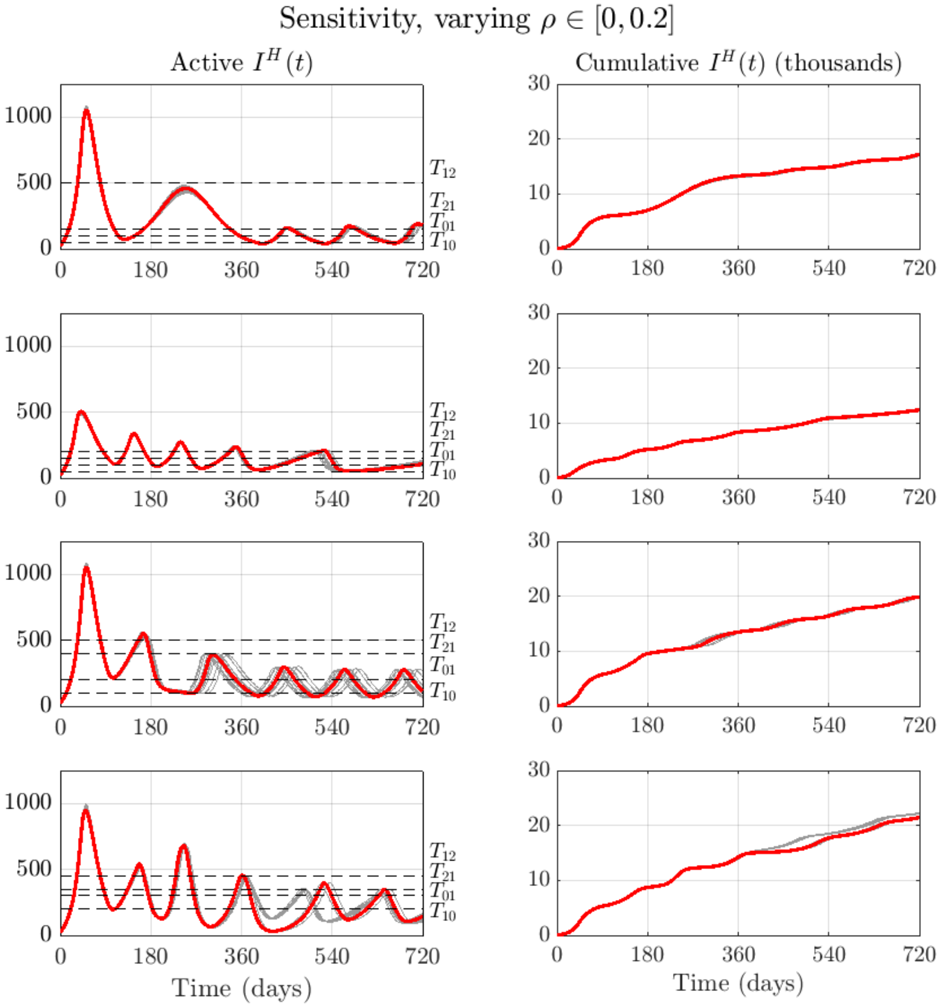


**Figure S12: Sensitivity to biological parameters.** Sensitivity in simulation dynamics to varying select parameters $\rho$ while keeping the remaining parameters fixed.

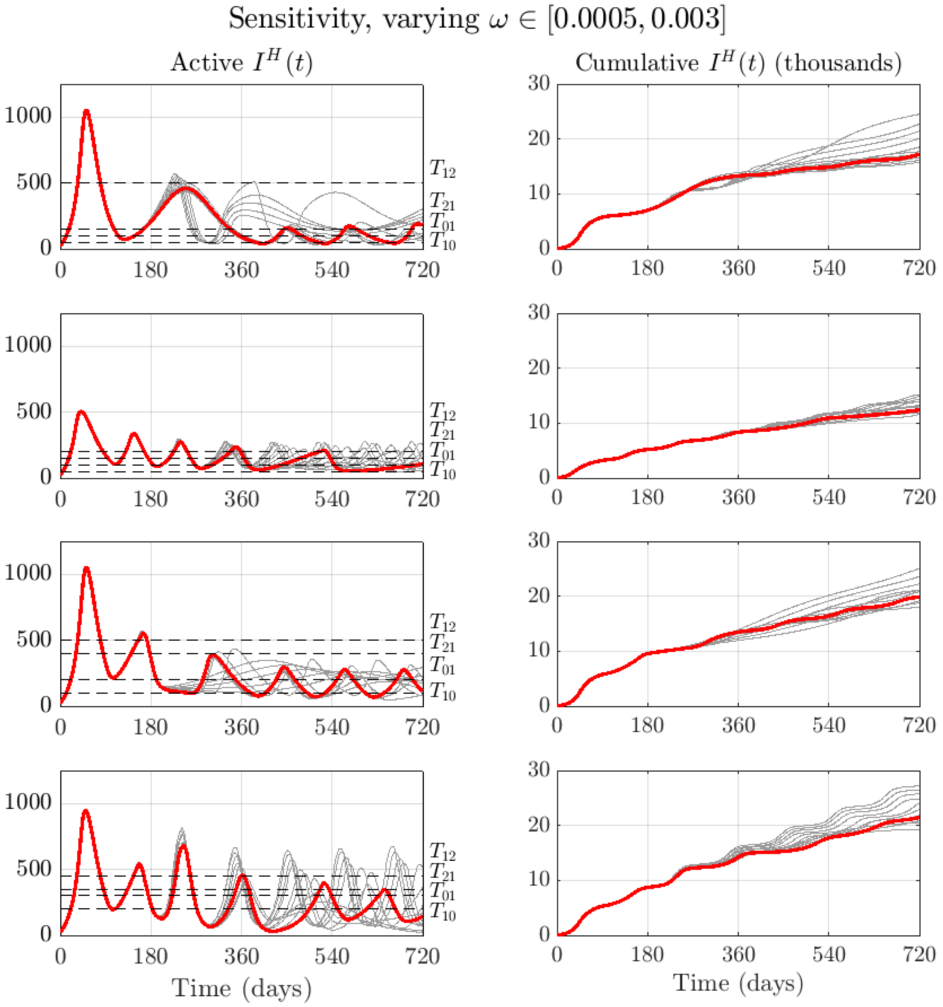


**Figure S13: Sensitivity to biological parameters.** Sensitivity in simulation dynamics to varying select parameters $\omega$ while keeping the remaining parameters fixed.

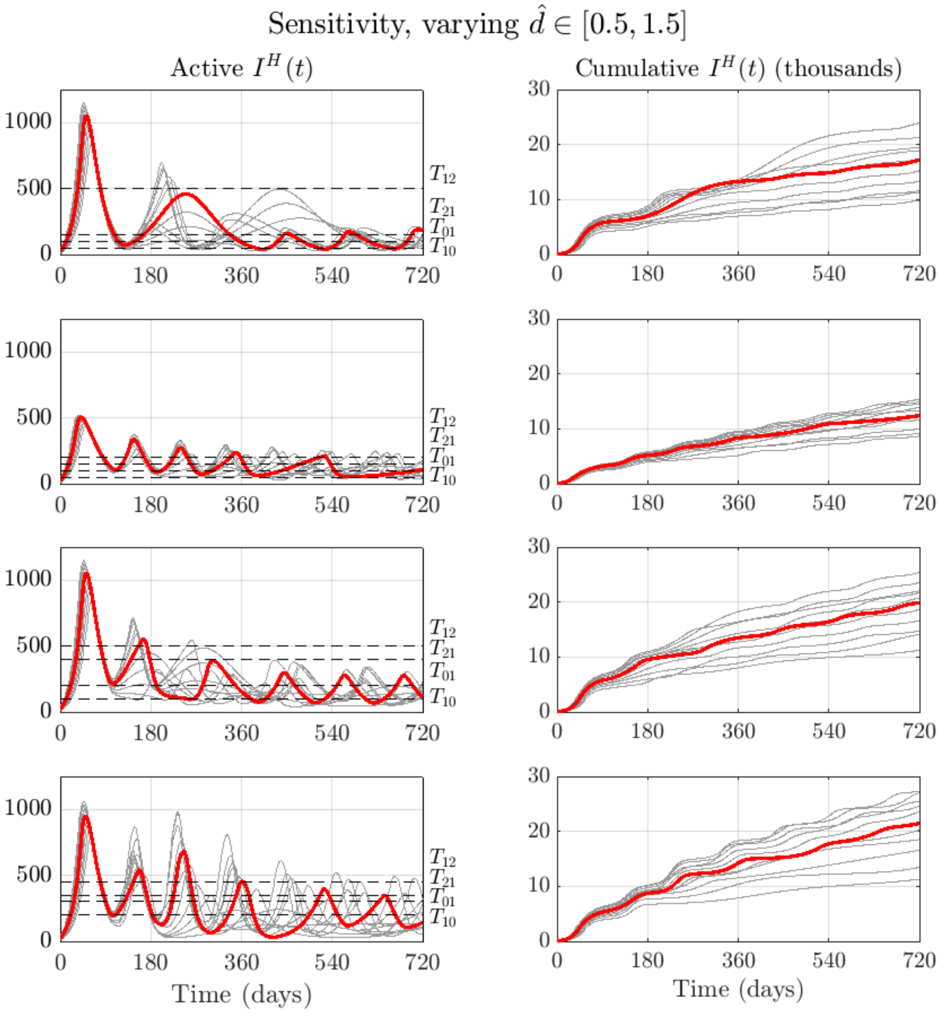


**Figure S14: Sensitivity to biological parameters.** Sensitivity in simulation dynamics to varying select parameters $d_a$ while keeping the remaining parameters fixed. Note $\hat{d}$ scales all values of $d_a$.

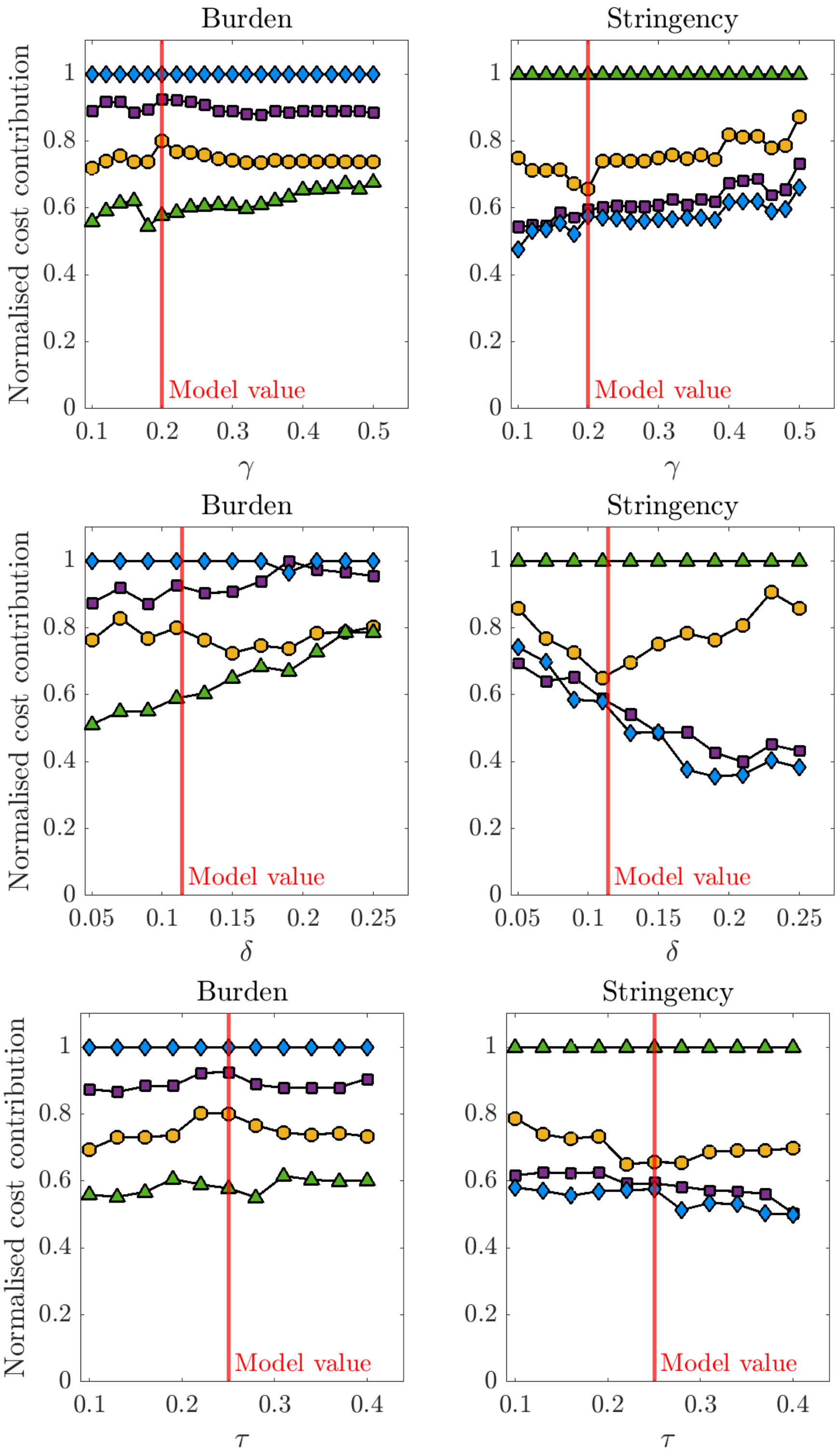


**Figure S15: Sensitivity to biological parameters.** Sensitivity in strategy cost contributions to varying select parameters ($\gamma$, $\delta$, $\tau$) while keeping the remaining parameters fixed. Costs are normalised against the worst strategy outcome, and its absolute value differs for each parameter value selection.

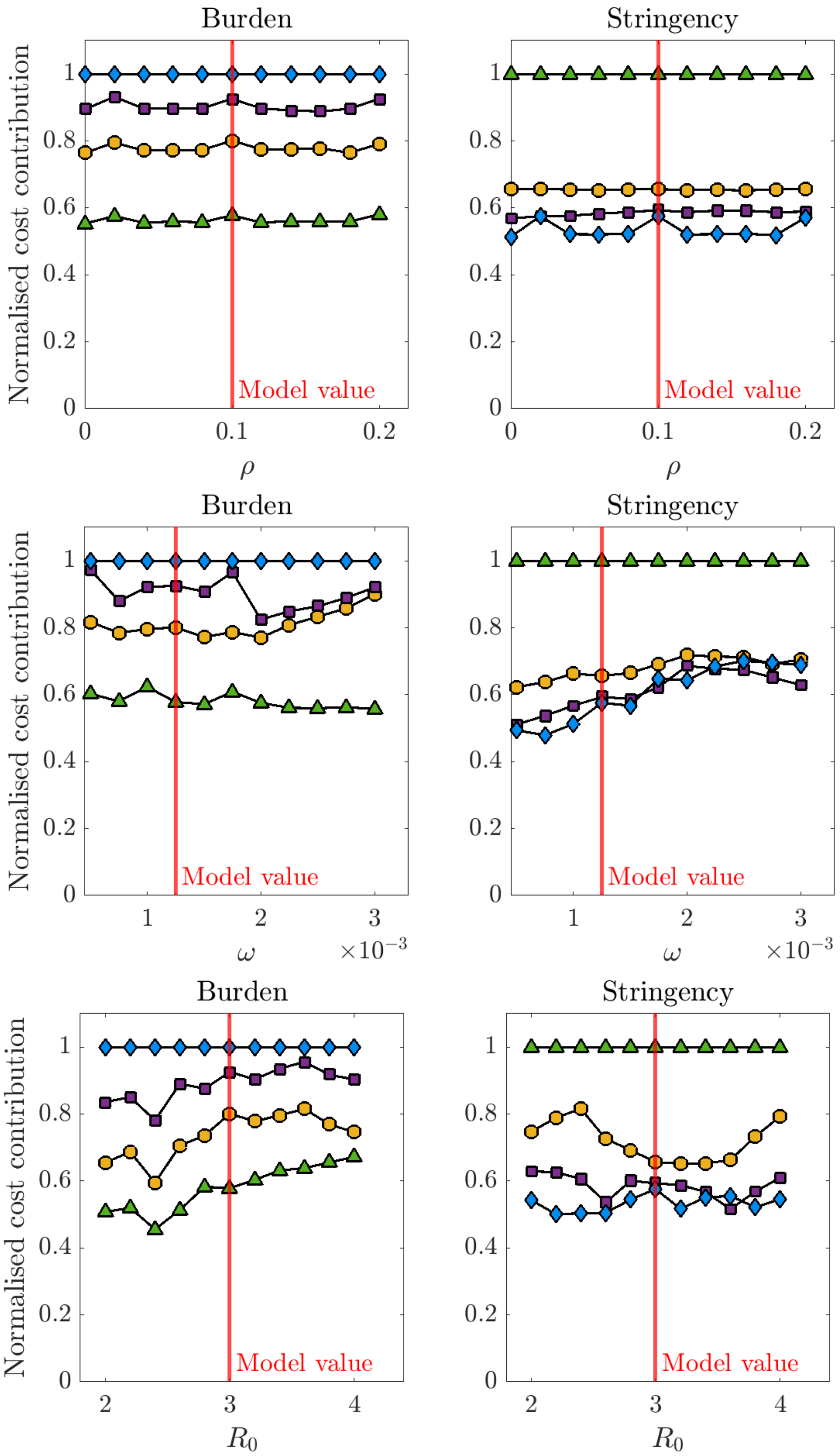


**Figure S16: Sensitivity to biological parameters.** Sensitivity in strategy cost contributions to varying select parameters ($\rho$, $\omega$, $R_0$) while keeping the remaining parameters fixed. Costs are normalised against the worst strategy outcome, and its absolute value differs for each parameter value selection.

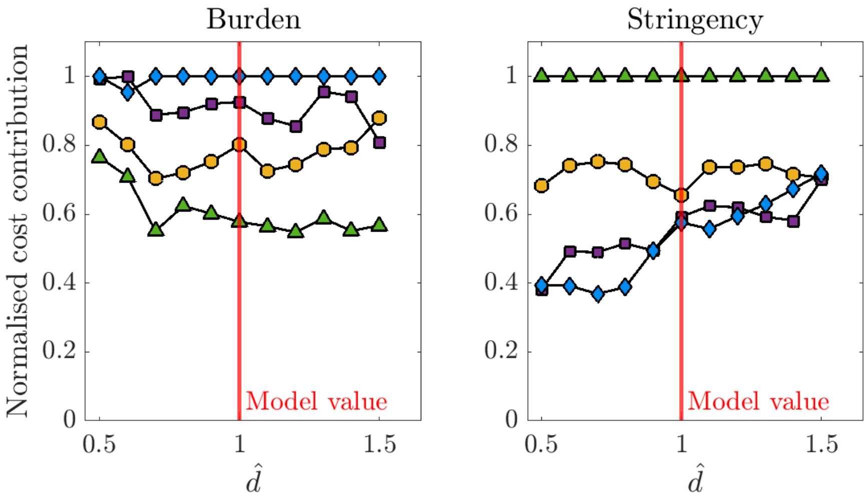


**Figure S17: Sensitivity to biological parameters.** Sensitivity in strategy cost contributions to varying select parameter $d_a$ while keeping the remaining parameters fixed. Costs are normalised against the worst strategy outcome, and its absolute value differs for each parameter value selection. Note $\hat{d}$ scales all values of $d_a$.

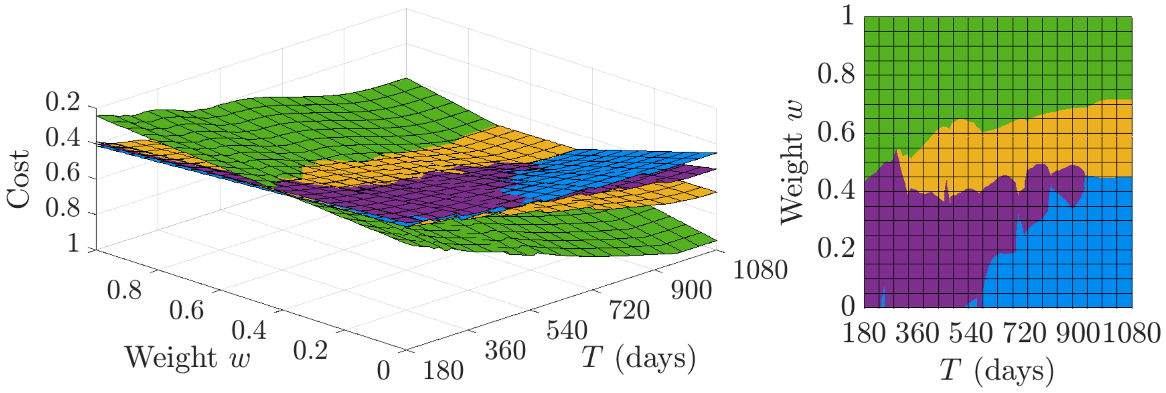


**(a)** $\eta = 0.9$, no discounting

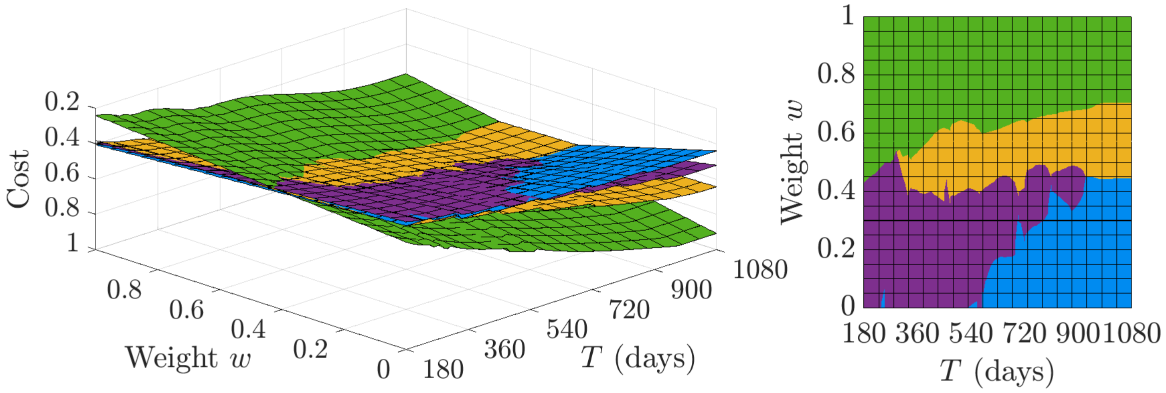


**(b)** $\eta = 0.9$, discounting 3.5% per annum

**Figure S18: Sensitivity to annual discounting.** Surface plots of cost (flipped z-axis) evaluated by the objective function (1) across weighting $w$ and different simulations which vary the arrival date of vaccination (left), and the top surface akin to viewing the optimal strategy for choice of $w$ and vaccine arrival date (right). Simulations were separately generated for vaccine eventual coverage $\eta = 0.9$, where we apply no discounting (a, main text results) and discounting of health and control costs at a per-annum rate of 3.5% (b). The control strategies are coloured as follows: yellow (Cautious easing), green (Suppression), purple (Slow control) and blue (Rapid control).
